# Menstrual cycle changes increased following COVID-19 mRNA vaccination: Social media validation and self-controlled case series analysis

**DOI:** 10.1101/2023.10.26.23297643

**Authors:** Aishwarya N Shetty, Gonzalo Sepulveda Kattan, Muhammad Javed, Christopher Pearce, Hazel J Clothier, Jim P Buttery

**Author notes:** **Correspondence to:** A/Prof Hazel J Clothier, 50 Flemington Road, Parkville, Australia 3052.

## Abstract

**Objectives:** To investigate if there was an increase in menstrual abnormality related presentation post COVID-19 vaccination.

**Design:** BERTopic machine learning, with a guided topic modelling option was used to analyse mentions of menstrual change in relation to COVID-19 vaccination on the social media platform Reddit. Self-controlled case series (SCCS) analysis using general practice data collected via the POpulation Level Analysis and Reporting (POLAR) tool with permission from Primary Health Networks (PHNs) as the de-identified dataset owners in Victoria and New South Wales.

**Setting:** Globally for social media analysis. Victoria and New South Wales (NSW), Australia for POLAR

**Participants:** For social media analysis, people who made a Reddit post about menstrual concerns post COVID-19 vaccine. For the SCCS analysis, people who presented to a POLAR GP registered practice with a new menstrual abnormality diagnosis.

**Exposures:** COVID-19 vaccination with adenovirus vector [AstraZeneca’s Vaxzervria® ChadOx1-S], mRNA [Pfizer-BioNTech’s Comirnaty® BNT162b2 and Moderna’s Spikevax®] or protein-subunit [Novavax’s Nuvaxovid®]).

**Outcomes and Measures:** Scraped social media posts were pre-processed, analysed for positive, negative, and neutral sentiments and topic modelled. Menstrual abnormality presentations of interest were isolated from the general practice dataset aggregated by POLAR, by searching for relevant SNOMED CT codes. Similarly, relative incidence (RI) was calculated for all COVID-19 vaccine types.

**Results:** Social media analysis saw peaks in menstrual change posts on Reddit since the global COVID-19 vaccine rollout. The SCCS analysis demonstrates an increase in general practice presentations of menstrual abnormality diagnosis following mRNA vaccines (RI= 1.14, 95% CI: 1.07 to 1.22, *P* <0.001).

**Conclusions and Relevance:** This study demonstrates an increase in menstrual abnormality presentations following COVID-19 mRNA vaccination. Our findings validate the concerns raised on social media so people who are vaccinated or are considering future vaccines feel heard, supported, and validated. Our analysis highlights the importance of using large real-world datasets to gather reliable evidence for public health decision making.

**Summary box:** Section 1: What is already known on this topic?

- Surveys and spontaneous surveillance systems suggested and association of menstrual cycle changes with COVID-19 vaccination.
- Heavy menstrual bleeding was added to the product information for mRNA vaccines in the European Union

Section 2: What this study adds?

- Our study is the first to prove an increase in menstrual abnormality related presentations post mRNA COVID-19 vaccines using routinely collected general practice data.
- Our findings validate the concerns raised by people who menstruate and help them with their future decision to vaccinate.

## Introduction

Since global COVID-19 vaccination began in 2021, community concerns of menstrual cycle changes following vaccination proliferated^1^. Social media reports described people who menstruate being left feeling “confused”, “fearful” and “dejected” as clinicians expressed scepticism about their concerns regarding a link with vaccination with no available evidence to confirm an association.^2^

Menstrual health is a sign of female health and is important to improving global population health^3, 4^.While variability is common, surveys, menstrual tracking apps and spontaneous surveillance systems added to growing data suggesting an association of menstrual cycle changes with COVID-19 vaccination.^5^ A 2023 cohort study did not find an association between increased menstrual cycle disorder consultations and COVID-19 vaccination in pre-menopausal women^3^.Heavy menstrual bleeding was added to the product information for mRNA vaccines in the European Union in October 2022^6^.

In the “infodemic” era where misinformation is rampant and can fuel vaccine hesitancy it is imperative that we can effectively listen and robustly investigate community vaccine safety concerns^7^. To ensure that the concerns regarding menstrual health were heard and appropriate analysis conducted to validate community concerns we 1) interrogated social media to explore the trends in COVID-19 vaccine related menstrual cycle change discussions and 2) analysed a large Australian primary care dataset to investigate if menstrual abnormality related consultations were more common within 6 weeks of any COVID-19 vaccination.

## Methods

We adopted a sequential approach where we assessed menstrual health concerns expressed on social media and then validated these concerns by conducting a self-controlled case series analysis on a large Australian general practice dataset. (Figure 1)

**Figure 1:**
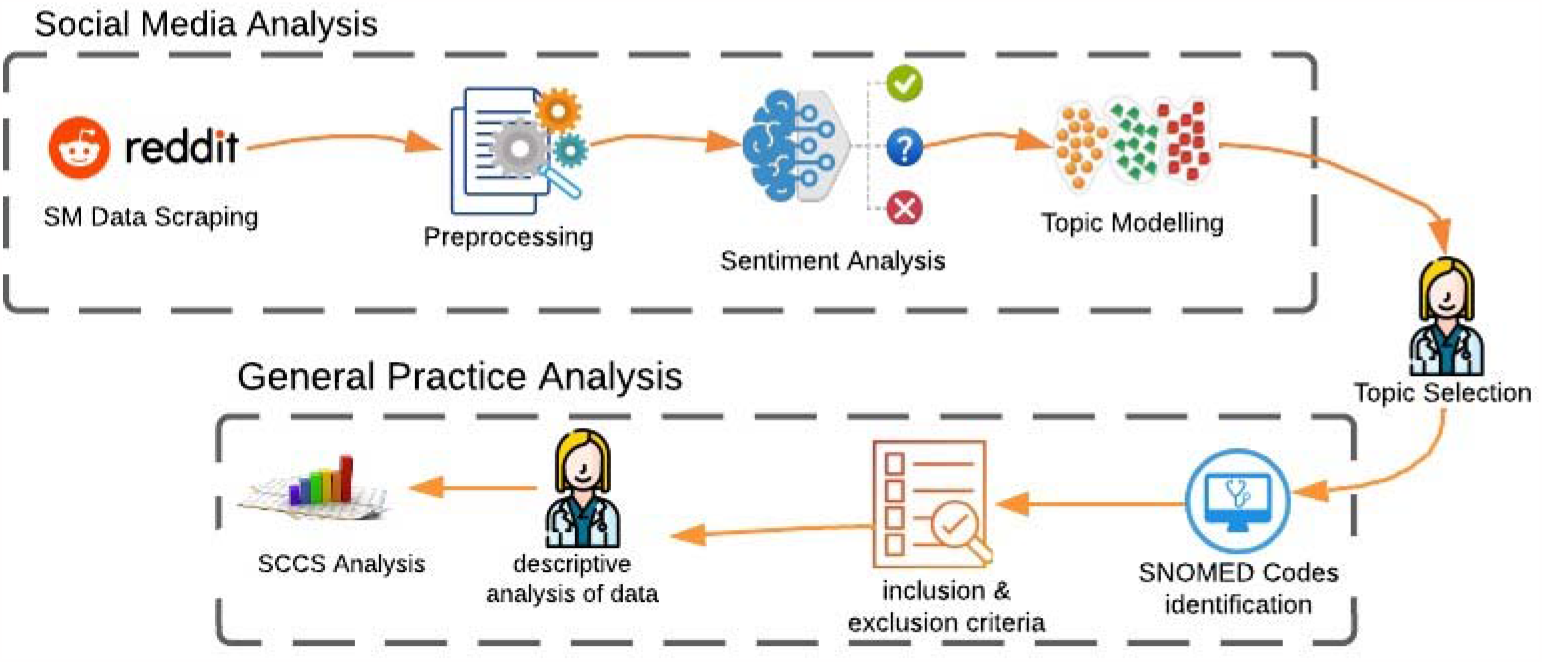
Sequential analysis approach

### Social media Analysis

We used BERTopic machine learning, with a guided topic modelling option, to analyse mentions of menstrual change with COVID-19 vaccination on the social media platform Reddit (Figure 1). Reddit is a global online community where registered users can submit content, engage in discussions, and interact with others on a wide range of topics^8^. Reddit social media posts were identified in a *data scraping phase, using* PRAW, a Python wrapper for the Reddit Application Performing Interface (API)^9^, to extract English posts. This was followed by a *pre-processing phase* where we removed hyperlinks and special characters. We converted any text contractions to long formats, split concatenated words, and converted emojis to text. Next the *sentiment analysis* phase was implemented which classifies the emotions and opinions in the posts as positive, negative, or neutral. We finetuned the COVID Twitter-BERT V2 (CT-BERT V2) model^10^, using manually labelled posts. Posts that showed encouraging attitudes towards COVID vaccines despite the news of AEFI were labelled as positive, while tweets showing discouraging reactions or refusal to take the vaccine were labelled negative. Tweets where users expressed neither sentiment in relation to AEFI, were marked as neutral^11^.

Finally in the *topic modelling* phase we only processed the posts categorised as negative and neutral for topic modelling, because these posts were likely to discuss vaccine hesitancy. Topic modelling helps understand the topics discussed in the sentiments which assists with understanding the opinion and topics of discussion of the public^11^.

### Self-controlled case series (SCCS) Analysis

We analysed a de-identified primary care dataset—Outcome Health’s Population Level Analysis and Reporting (POLAR) deployed in partnership with primary health networks in the Australian states of Victoria and New South Wales namely: Central and Eastern Sydney PHN, Eastern Melbourne PHN, Gippsland PHN, South Eastern Melbourne PHN, and South Western Sydney PHN. POLAR provides near real-time routinely collected electronic general practice (GP) health information representing a pooled catchment of 12 million patients.^4^ De-identified data were accessed to identify GP consultations of people categorised as female in the dataset, aged 15–49 years with a new menstrual cycle change between 1 January 2021 and 28 March 2023. A menstrual abnormality related diagnosis included any premenstrual or menstruation disruptions, change in flow and/or cycle irregularities (supplementary Table 1) identified using SNOMED CT codes^12^. Using a SCCS design^13^, we calculated the relative incidence of menstrual cycle change within a 42-day risk window post any COVID-19 vaccination and by COVID-19 vaccine type (adenovirus vector [Vaxzervria® ChadOx1-S, AstraZeneca], mRNA [Comirnaty® BNT162b2, Pfizer-BioNTech and Spikevax®, Moderna] or protein-subunit [Nuvaxovid®, Novavax]). All other time periods outside the 42-day risk window and within the observation period constituted the baseline period. (Supplementary Figure 1) Analysis was performed in R (version 4.2.3)^14^, P values were considered significant at <0.05.

#### Patient and Public Involvement

Public sentiments were heard using our social media analysis, which was then validated using the POLAR SCCS analysis. The POLAR data were completely de-identified and hence it was not possible to involve patients.

## Results

Overall, 70,355 social media posts were retrieved, which after pre-processing resulted in 65,254 posts. Sentiment analysis step demonstrated a majority neutral posts (64.5%, n= 42,106), followed by positive (25.48%, n=16624) and negative posts (10%, n=6524) (Figure 2). Some manually labelled sample sentiments are listed in Table 1.

**Figure 2.**
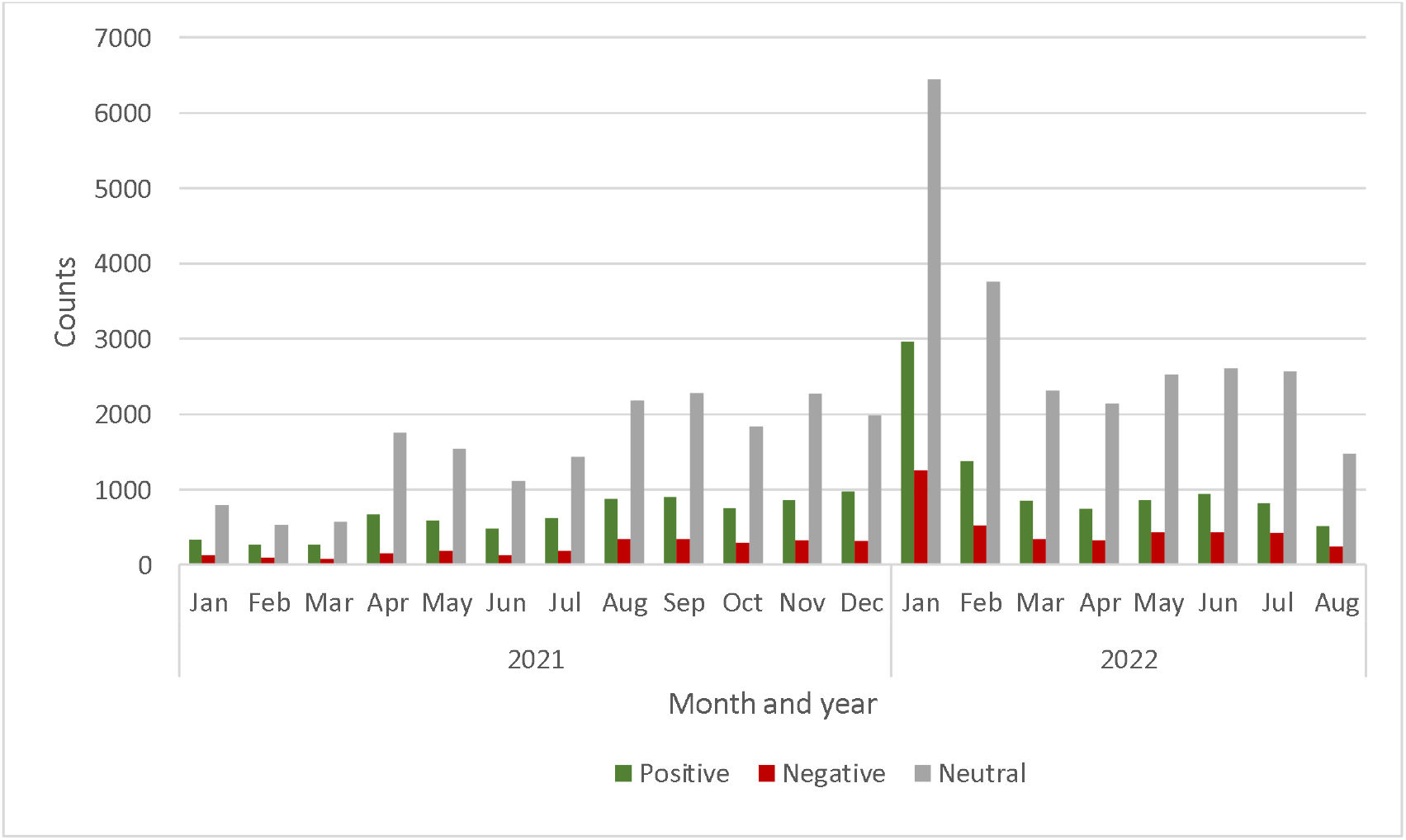
Monthly distribution of positive, negative, and neutral sentiments

**Table 1:** Sample negative and neutral sentiments.

| Polarity | Posts |
| --- | --- |
| Negative | “I got the Moderna vaccines a couple weeks ago. a week later I had by far the worst menstrual cycle in my entire life. it put me down for three days.”<br>“As a woman it is extremely disturbing the amount of women on that sub that are having menstrual side effects.”<br>“The way they swept women s menstrual changes under the rug is criminal. i think the last report said something like 50 of women have reported a change in their menses after the vaccine. and they are just acting like it is no biggie.” |
| Neutral | “Someone posted this in a different thread since covid has an affect on menstrual cycles perhaps the vaccine has an affect as well?”<br>“Has there been anything published to show these vaccines have any effect on menstrual cycles?”<br>“First one did not impact my period and I was on it when i got it. second one triggered mine to come 4 days early but it impacted it weird. it was like a 4 day teaser than back to normal flow cycles. I chalked it up to my body fighting off stuff like it was sick and I know sickness and stress can really muck with mine” |

Topic modelling demonstrated a women and children’s hesitancy theme in which menstrual cycle was the most frequently discussed topic (2890 posts). The distribution of these menstrual cycle posts saw two prominent peaks; the first peak around April-May 2021(350-400 posts) and in January 2022(389 posts) which is likely due to the commencement of the global COVID-19 vaccine rollout^15^ and twitter post from a social media influencer on the topic^16^. (Figure 3)

**Figure 3:**
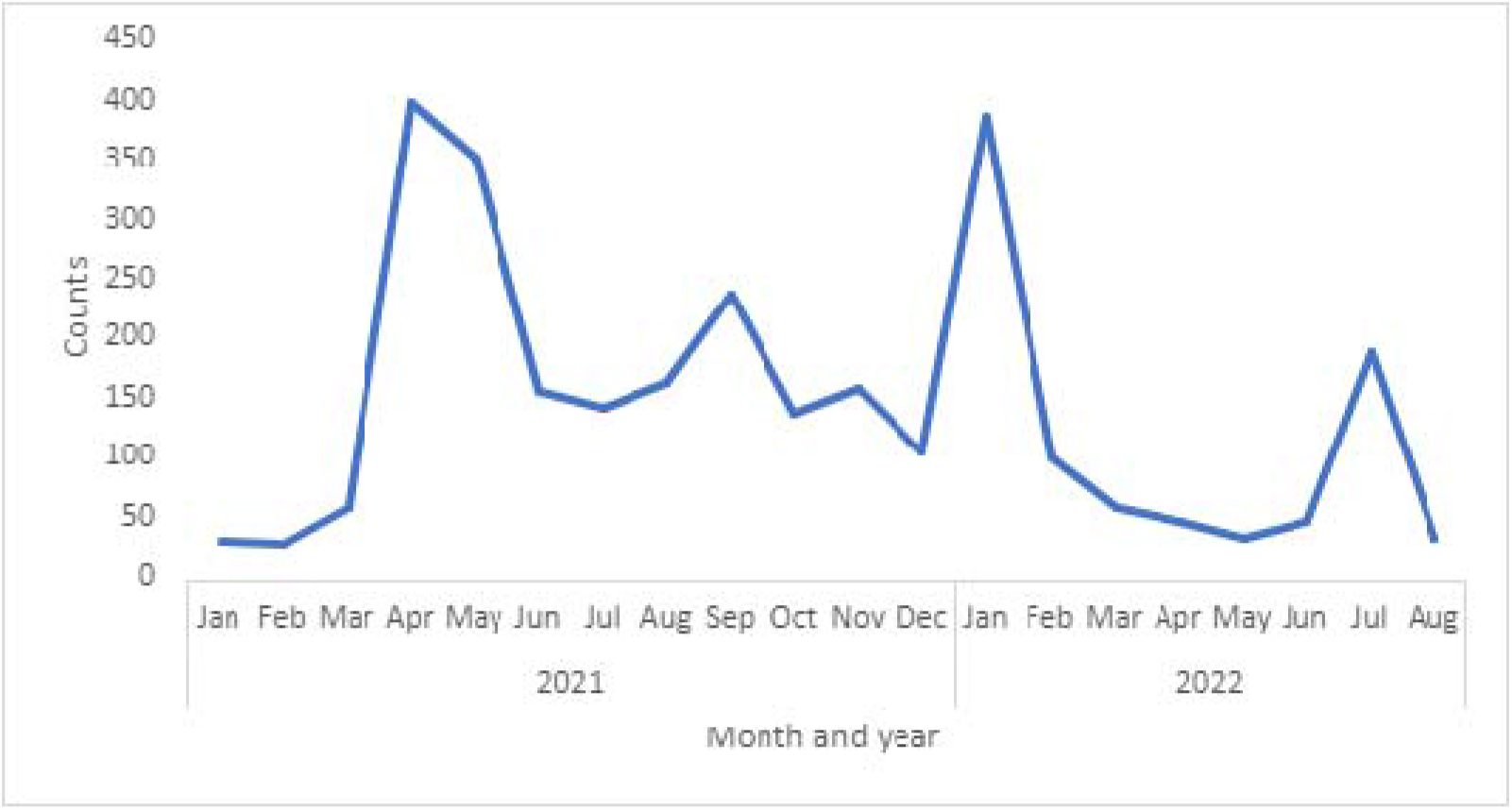
Monthly distributions of neutral and negative menstrual cycle posts on Reddit

### SCCS analysis

During the study period, 40,750 people categorised as females had 46,353 presentations with menstrual cycle changes. Of these, 22,145 had received COVID-19 vaccination (Comirnaty®=19,392, Spikevax®=511, Vaxzevria®=2,176, Nuvaxovid®=56, unknown=10) and 1,063 had a new menstrual cycle change within 42-days of vaccination.

The incidence of a new menstrual cycle change presentation was higher in the 42-day post-vaccination period (relative incidence 1.15, 95%CI:1.08 to 1.22, P<.001) than the baseline period. When analyzed by vaccine type, only mRNA vaccines (Comirnaty® and Spikevax®) were associated with the higher incidence of menstrual cycle change presentation (Table 2). On extending the risk window beyond 42 days, an increase in presentation was still evident at 49 days (Relative incidence 1.32, 95%CI:1.13 to 1.53, P=<0.001) but dissipated by 90 days post vaccination. (Relative incidence 1.11, 95%CI:0.94 to 1.32, P=0.20)

**Table 2:** Menstrual cycle change presentations within 42-days post COVID-19 vaccination by vaccine type.

| Vaccine type* | Relative incidence | 95% Confidence interval | p-value |
| --- | --- | --- | --- |
| mRNA (Comirnaty® and Spikevax®) | 1.14 | 1.07 to 1.22 | <0.001 |
| Adenoviral-vectored (Vaxzevria®) | 1.15 | 0.94 to 1.40 | 0.15 |
| Protein-based (Nuvaxovid®) | 1.41 | 0.50 to 3.93 | 0.52 |

\*COVID-19 vaccine brands licensed for use in Australia<sup>15</sup>

## Discussion

Our social media analysis demonstrated that people expressed concerns regarding changes to their menstrual cycle after receiving the COVID-19 vaccination. This was validated by our self-controlled case series analysis of a large primary care dataset analysis demonstrating that mRNA COVID-19 vaccination is associated with a higher incidence of new menstrual cycle change in the 6 weeks following vaccination. While a Swedish nationwide cohort study found no evidence of increased healthcare contacts in premenopausal women, our study included a longer duration, larger event outcome size, with the SCCS design controlling for fixed confounders.^13, 17^ In contrast, a Norwegian study observed an increased occurrence of menstrual changes post vaccination compared to the last cycle prior to vaccination, however, this study used a self-administered questionnaire which is prone to recall bias and was conducted when media attention to menstrual changes was high possibly leading to an over estimation of association and was only conducted in women between 18-30 years of age^18^. Furthermore, a retrospective survey conducted in United Kingdom found that 18% of the pre-menopausal participants reported a menstrual change post their first dose of COVID-19 vaccine, however, it was not set up initially to specifically answer menstrual changes post COVID-19 vaccination (secondary analysis) and was prone to reporter, selection and recall bias.^19^ Another study conducted on a menstrual tracking app in the U.S. found a temporary less than 1-day change in menstrual cycle length, however the findings of this study were not generalisable to the general population as people using the app may have certain common characteristics.^20^

In today’s era, social media is accessible to almost everyone, making it a valuable tool for public health organisations and consumers to connect and collaborate^21^. It removes the barrier of seeking medical attention or submitting a report, and therefore a useful adjunct source of health information for vaccine safety surveillance and early signal detection^22, 23^.

Accessing de-identified routinely collected health record data like POLAR for secondary analysis is recognised as an important adjunct to public health surveillance and removes the biases (reporting and recall) associated with most studies published in literature. Additionally, it is likely to be a more accurate representation of the true state of menstrual cycle changes in the community. Furthermore, the SCCS design is an established pharmacovigilance methodology to determine associations between discrete exposures such as vaccines, and subsequent health outcomes^24^.

### Limitations

Our social media analysis was restricted to English language only and the manual labelling of post could have introduced bias. It is possible not all exposures (vaccines) or outcomes (consultations) were included in the POLAR dataset resulting in a potential under-ascertainment of the associations we detected between vaccination and menstrual cycle disorder consultations.

## Conclusion

Our findings validate concerns expressed in social media and surveys that mRNA COVID vaccines were associated with changes in menstruation. These underline the importance of listening to community-level concerns about public health interventions including vaccines.^5^Coupled with existing information that these changes are temporary;^5, 17^ we trust this will validate the experiences and reassure those who have experienced menstrual cycle changes following vaccination. Knowledge of our findings will prepare people considering future vaccinations for temporarily altered cycles and make informed decisions. It will also help healthcare professionals counsel those who experienced menstrual change post COVID-19 vaccination and appropriately validate their concerns.

## Data Availability

No additional data available.

## Footnotes

### Ethical approval

This study was approved by Monash Human Research Ethics Committee [RES-18-0000-232A].

## Acknowledgements

We thank Mr. Adam McLeod and Karina Gardner of Outcome Health, Melbourne, Victoria, for their support with this research and Dr Diana Vlasenko (Health Informatics, Centre for Health Analytics, Melbourne Children’s Campus, Victoria) for assistance with mapping of SNOMED CT codes. They did not receive any compensation for these roles.

## Contribution to Authorship

Shetty, Kattan had access to POLAR data and Javed the social media data and take responsibility for the integrity of the data and the accuracy of the data analysis.

Concept and design: Shetty, Buttery, Clothier

Acquisition, analysis, or interpretation of data: All authors.

Drafting of the manuscript: Shetty, Javed, Clothier

Critical revision of the manuscript for important intellectual content: All authors.

Statistical analysis: Shetty, Kattan, Javed, Clothier

Administrative, technical, or material support: Shetty

## Transparency declaration

The lead author (AS) affirm that the manuscript is an honest, accurate, and transparent account of the study being reported; that no important aspects of the study have been omitted; and that any discrepancies from the study as planned (and, if relevant, registered) have been explained.

## Dissemination statement

Dissemination to participants and related patient and public communities: The research findings will be disseminated to the wider community through press releases, social media platforms, presentations at international forums, and reports to relevant government agencies and academic societies.

## References

1. Katz A, Tepper Y, Birk O, Eran A. Web and social media searches highlight menstrual irregularities as a global concern in COVID-19 vaccinations. Scientific Reports. 2022;12(1):17657.

2. Liew TM, Lee CS. Examining the Utility of Social Media in COVID-19 Vaccination: Unsupervised Learning of 672,133 Twitter Posts. JMIR Public Health Surveill. 2021;7(11):e29789.

3. Hennegan J, Winkler IT, Bobel C, Keiser D, Hampton J, Larsson G, et al. Menstrual health: a definition for policy, practice, and research. Sexual and Reproductive Health Matters. 2021;29(1):31–8.

4. United Nations. Pushing menstrual health on the 2030 Agenda New York2018 [cited 2023. Available from: https://sdgs.un.org/events/pushing-menstrual-health-2030-agenda-28082.

5. Chao MJ, Menon C, Elgendi M. Effect of COVID-19 vaccination on the menstrual cycle. Front Med (Lausanne). 2022;9:1065421.

6. European Medicines Agency. COVID-19 vaccines safety update 2022 [Available from: https://www.ema.europa.eu/en/documents/covid-19-vaccine-safety-update/covid-19-vaccines-safety-update-10-november-2022_en.pdf.

7. Clothier HJ, Crawford NW, Russell M, Kelly H, Buttery JP. Evaluation of ‘SAEFVIC’, a pharmacovigilance surveillance scheme for the spontaneous reporting of adverse events following immunisation in Victoria, Australia. Drug safety. 2017;40:483–95.

8. Reddit Inc. Dive Into Anything [Available from: https://www.redditinc.com/.

9. PRAW. PRAW: The Python Reddit API Wrapper [Available from: https://praw.readthedocs.io/en/stable/.

10. Müller M, Salathé M, Kummervold PE. Covid-twitter-bert: A natural language processing model to analyse covid-19 content on twitter. Frontiers in Artificial Intelligence. 2023;6:1023281.

11. Javed M BJ, Khademi S, Hickman J, Clothier HJ, Palmer C, Dimaguila GL. VaxPulse: Monitoring of Online Public Concerns to Enhance Post-licensure Vaccine Surveillance: Rationale and Study Design[Preprint] 2023.

12. Donnelly K. SNOMED-CT: The advanced terminology and coding system for eHealth. Studies in health technology and informatics. 2006;121:279.

13. Petersen I, Douglas I, Whitaker H. Self controlled case series methods: an alternative to standard epidemiological study designs. bmj. 2016;354.

14. Team RDC. R: A language and environment for statistical computing. (No Title). 2010.

15. TGA. COVID-19 vaccines regulatory status 2023 [Available from: https://www.tga.gov.au/products/covid-19/covid-19-vaccines/covid-19-vaccines-regulatory-status.

16. Jones M. Social Media Influencers Are Spreading Wild Rumors About COVID-19 Vaccines and Periods: 2021 [Available from: https://www.motherjones.com/politics/2021/04/social-media-influencers-are-spreading-wild-rumors-about-covid-19-vaccines-and-periods/.

17. Ljung R, Xu Y, Sundström A, Leach S, Hallberg E, Bygdell M, et al. Association between SARS-CoV-2 vaccination and healthcare contacts for menstrual disturbance and bleeding in women before and after menopause: nationwide, register based cohort study. bmj. 2023;381.

18. Trogstad L, Laake I, Robertson AH, Mjaaland S, Caspersen IH, Juvet LK, et al. Heavy bleeding and other menstrual disturbances in young women after COVID-19 vaccination. Vaccine. 2023;41(36):5271–82.

19. Alvergne A, Kountourides G, Argentieri MA, Agyen L, Rogers N, Knight D, et al. A retrospective case-control study on menstrual cycle changes following COVID-19 vaccination and disease. iScience. 2023;26(4):106401.

20. Edelman A, Boniface ER, Benhar E, Han L, Matteson KA, Favaro C, et al. Association Between Menstrual Cycle Length and Coronavirus Disease 2019 (COVID-19) Vaccination: A U.S. Cohort. Obstet Gynecol. 2022;139(4):481–9.

21. Chen J, Wang Y. Social media use for health purposes: systematic review. Journal of medical Internet research. 2021;23(5):e17917.

22. Buttery JP, Clothier H. Information systems for vaccine safety surveillance. Human Vaccines & Immunotherapeutics. 2022;18(6):2100173.

23. Clothier HJ, Lawrie J, Russell MA, Kelly H, Buttery JP. Early signal detection of adverse events following influenza vaccination using proportional reporting ratio, Victoria, Australia. Plos one. 2019;14(11):e0224702.

24. Joy M, Agrawal U, Fan X, Robertson C, Anand SN, Ordonez-Mena J, et al. Thrombocytopenic, thromboembolic and haemorrhagic events following second dose with BNT162b2 and ChAdOx1: self-controlled case series analysis of the English national sentinel cohort. The Lancet Regional Health–Europe. 2023;32.

