## Supplementary tables for "Menstrual cycle changes increased following COVID-19 mRNA vaccination: Social media validation and self-controlled case series analysis"

Supplementary Table 1: SNOMED CT text and codes included in the menstrual cycle changes SCCS analysis.

| **SNOMED CT text** | **SNOMED CT code** |
| --- | --- |
| Oligomenorrhoea | 52073004 |
| Amenorrhoea | 14302001 |
| Menstrual problem | 289896006 |
| Premenstrual tension syndrome | 82639001 |
| Suppression of menstruation | 198425001 |
| Irregular periods | 80182007 |
| Menorrhagia | 386692008 |
| Intermenstrual bleeding - irregular | 64996003 |
| Abnormal vaginal bleeding | 301822002 |
| Dysmenorrhoea | 266599000 |

Supplementary Figure 1: Observation period and risk window applied in the SCCS model.

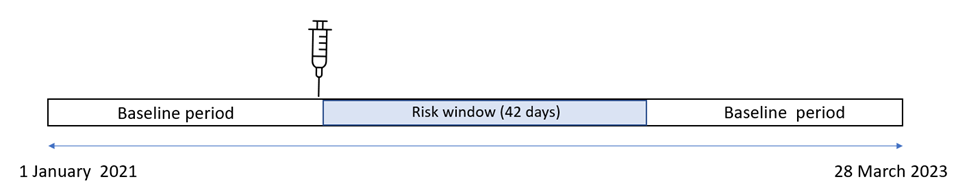
